# Multi-parametric disease dynamics study and analysis of the COVID-19 epidemic and implementation of population-wide intrusions: The Indian perspective

**DOI:** 10.1101/2020.06.02.20120360

**Authors:** Samuel Bharti, Priyanka Narad, Parul Chugh, Alakto Choudhury, Seema Bhatnagar, Abhishek Sengupta

**Affiliations:** Amity Institute of Biotechnology, Amity University, Uttar Pradesh, INDIA

**Keywords:** COVID-19, Indian outbreak, Population Density, Mobility, Forecasting, Crowdsourced database

## Abstract

The outbreak of COVID-19 had spread at a deadly rate since its onset at Wuhan, China and is now spread across 216 countries and has affected more than 6 million people all over the world. The global response throughout the world has been primarily the implementation of lockdown measures, testing and contact tracing to minimise the spread of the disease. The aim of the present study was to predict the COVID-19 prevalence and disease progression rate in Indian scenario in order to provide an analysis that can shed light on comprehending the trends of the outbreak and outline an impression of the epidemiological stage for each state of a diverse country like India. In addition, the forecast of COVID-19 incidence trends of these states can help take safety measures and policy design for this epidemic in the days to come. In order to achieve the same, we have utilized an approach where we test modelling choices of the spatially unambiguous kind, proposed by the wave of infections spreading from the initial slow progression to a higher curve. We have estimated the parameters of an individual state using factors like population density and mobility. The findings can also be used to strategize the testing and quarantine processes to manipulate the spread of the disease in the future. This is especially important for a country like India that has several limitations about healthcare infrastructure, diversity in socioeconomic status, high population density, housing conditions, health care coverage that can be important determinants for the overall impact of the pandemic. The results of our 5-phase model depict a projection of the state wise infections/disease over time. The model can generate live graphs as per the change in the data values as the values are automatically being fetched from the crowd-sourced database.

## Introduction

A Pneumonia like viral disease of an unknown etiology was reported to the World Health Organization (WHO) from Wuhan province, China on 31^st^ December 2019 [Sohrabi et al. 2020]. WHO on February 11^th^ 2020 announced that the new coronavirus disease was COVID-19. The highly contagious nature of the virus is not related to any of the previous outbreaks of SARS and MERS making COVID-19 as a deadly pandemic of the 21^st^ century [Sreeram, 2020]. COVID-19 was declared a public health emergency on 30^th^ January 2020. India reported its first COVID-19 case on the same day. As of 28th May 2020, the Ministry of Health and Family Welfare, Government of India has updated 158,333 cases, 67,692 recoveries (including 1 migration) and 4,531 deaths in the country [MOHFW, 2020]. Although, the surge of the disease outbreak was steady in the initial days of the spread in the Indian context, the Indian scenario with the progression of disease has been very different and several measures were taken to break the chain of transmission. Amongst the prominent public health measures taken included an early nationwide lockdown, the suspension of inbound International air travel, rigorous contact tracing and follow up with affected patients. India, however, is a geographically, climatically, and socially; a country of tremendous diversity with a population density of more than 1.3 billion [Sarma, 2015]. The population density varies significantly across major metro cities and smaller cities and there is a prominent divide in the urban and rural context [Pradhan, 2017]. Significantly, the population density in India is 464 people per km square which is indeed a matter of grave concern for the spread of a highly contagious pathogen such as COVID-19 [Economic Times, 2020]. COVID-19 has been compared to SARS-CoV and MERS-CoV in several papers in the past; however, the epidemiology of COVID-19 has been unusual and not related to any of the previous coronavirus outbreaks [ICMR, 2020]. Recent literature on COVID-19, indicate that it has a reproduction number (R_o_) of 2.0-3.5 [Park et al. 2020].

The trajectory of upward surge in the disease outbreak was evident from the available figures mentioned below. The number of cases rose from 1-20000 in 80 days from the first detected case, from 20000-40000 in 12 days, from 40000-80000 in 11 days and from 80000-100000 in 4 days. The numbers are a mounting concern for the people of India [Yahoo News, 2020], the nationwide lockdown has been extended in phases spread over a period of 60 days continuing in parts till date. On 28th May 2020, the Ministry of Health and Family Welfare, Government of India have updated 158,333 cases, 67,692 recoveries (including 1 migration) and 4,531 deaths. Further, India has conducted 2,404,267 tests which account to 1744 tests per million of the population [Our World in Data, 2020]. The current infection has a spread rate of 4.3% of the total tests coming positive among the total number being tested; approximately 0.007% of the total population has been–infected till date.

Predictive modeling can efficiently track and forecast the outbreak and future course of a pandemic [Wang et al. 2020]. In Indian context, there are broadly four major epidemiology models used for predicting the Covid-19 progression (Mahalle et al.2020). The first is a model developed by scientists at the Indian Council of Medical Research (ICMR) and their collaborators. The second is a model produced by a group of epidemiologists and statisticians from the University of Michigan (University of Michigan, 2020). The third is a set of reports published by the Centre for Disease Dynamics, Economics and Policy (CDDEP) at Johns Hopkins University. Finally, there is a recent study from scientists at Cambridge University, with one of the authors also affiliated with the Institute of Mathematical Sciences, Chennai.

The above models however ignore many features such as geographical density and population driven factors to name a few and consider India as a single unit which practically does not translate into a realistic model of the outbreak for the Indian scenario. These models also do not account for the mobility patterns such as movement to and from various parts of the country that has been a crucial factor in transmission and spread of disease in India specifically. This aspect is significant and should be accounted for while creating predictive models for a diverse country such as India.

## Results

The aim of the present study was to predict the COVID-19 prevalence and disease progression rate in Indian scenario in order to provide an analysis that can shed light on comprehending the trends of the outbreak and outline an impression of the epidemiological stage for each state of a diverse country like India. In addition, the forecast of COVID-19 prevalence trends of these states can help take safety measures and policy design for this epidemic in the days to come. In order to achieve the same, we have utilized an approach where we test modelling choices of the spatially unambiguous kind, proposed by the wave of infections spreading from the initial slow progression to a highly increasing curve. We estimate parameters of an individual state using factors like population density and mobility.

The demographic distribution of the disease as per reports from other countries indicates that the disease occurrence and fatality is more in males than in females [Salonia et al. 2020]. To corroborate this in the Indian context we estimated number of female and male ratio as well as the age group of an individual affected by COVID-19 as of 28^th^ May 2020 in India. Results of the first phase of our model found that 65.6% of affected individuals were male, whereas 34.6% were female (Figure 2A). As model categorizes age of affected individuals in 5-year age group, Figure 2B indicates the count of affected cases present in each group. Individuals belonging to the age groups i.e. 21-25, 26-30, 31-35 and 36-40 were observed to be most affected; with highest number of observed cases in 26-30 age group. Also, in the most vulnerable age groups it was significantly observed that females were less likely to be affected by the disease as compared to males (Fig 3). This observation was in line with the estimates worldwide [Wiemers et al. 2020].Moreover, being in the younger age group it is expected that most cases may be asymptomatic with lesser incidence of fatality [Verity et al. 2020]. This is in sharp contrast with countries such as Italy where nearly 69% of infected population was in the age group ranging between 51-70 years. Accordingly the highest fatality was also observed in this age group [The Wall Street Journal, 2020].

**Figure 2A & 2B:**
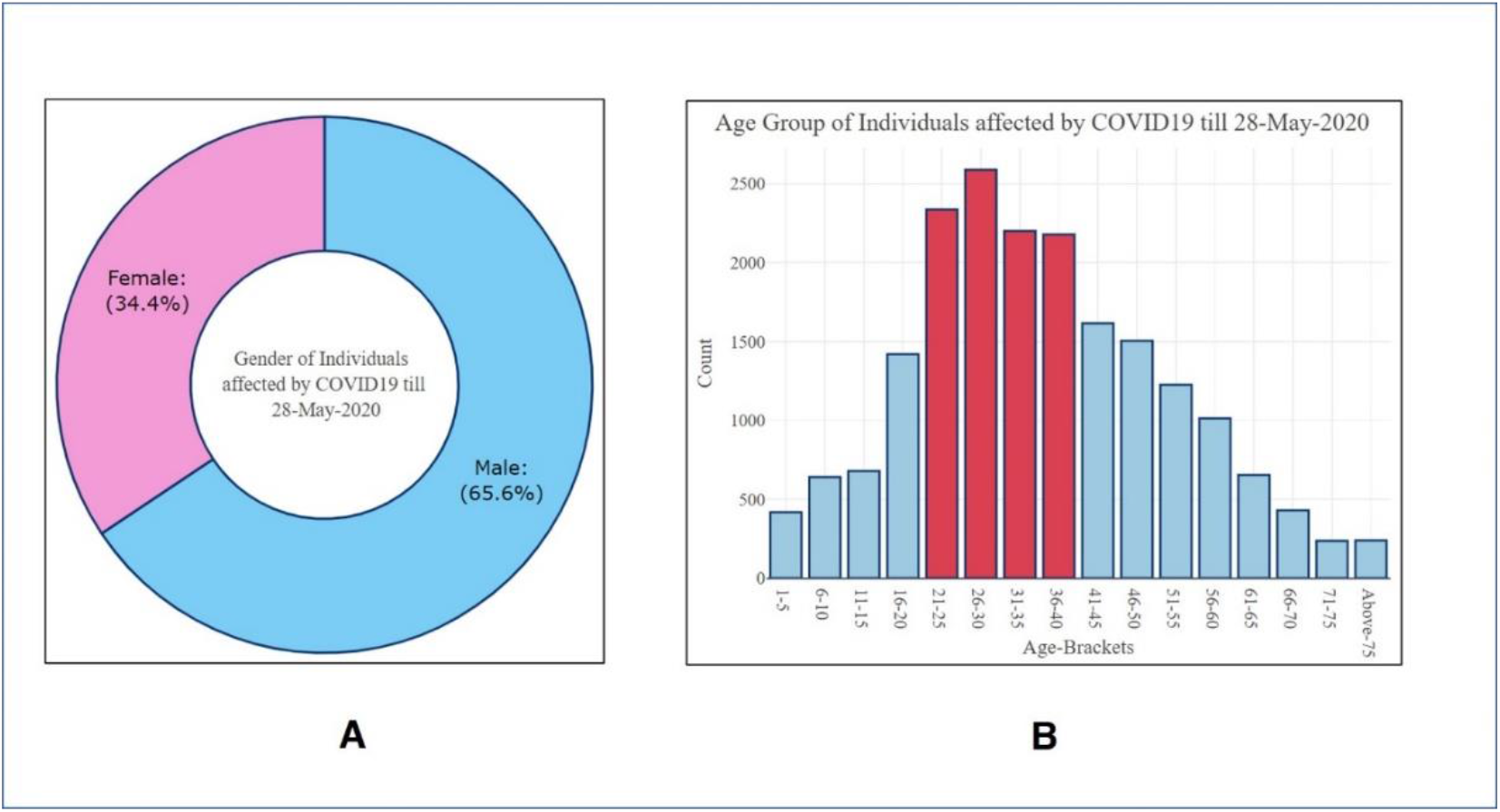
Representation of Gender and Age distribution of COVID-19 in India

**Figure 3:**
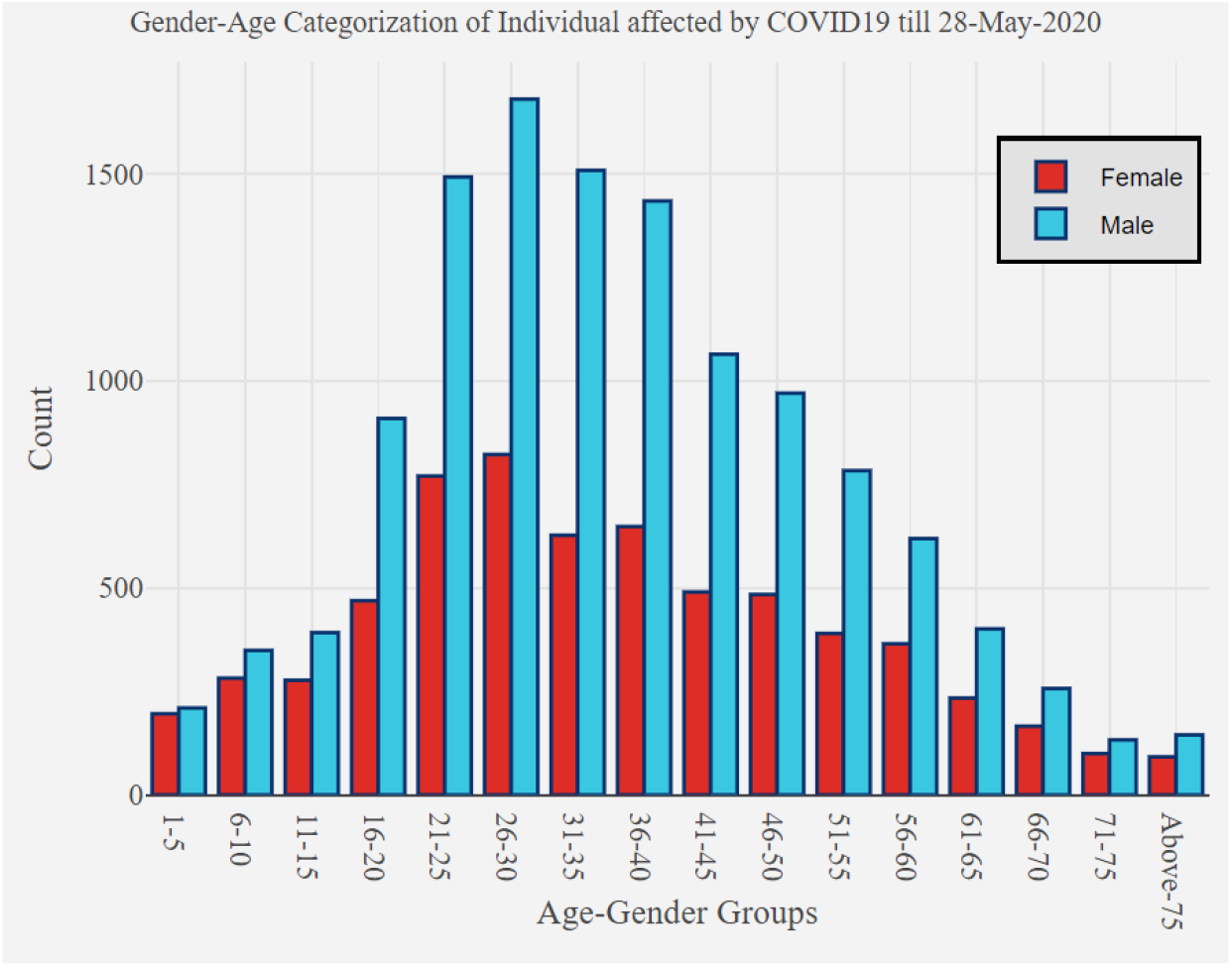
Gender Age Categorization of Individual affected by COVID-19

Further, we estimated beta and gamma values for 10 most affected states in India. Figure 4 depicts the estimated values for each state as well as the current calculated reproductive number (Rn1) till 28^th^ May 20. Maharashtra; which is currently leading with highest number of confirmed cases has calculated Rn1 of 1.023, followed by Tamil Nadu with Rn1 of 1.014, Delhi with Rn1 of 1.026. The beta and gamma values are important estimates to study the progression of disease whereby Beta is the infection rate of the pathogen, and gamma is the recovery rate. Together, these two values give the basic reproduction number Rn1: the average number of secondary infections caused by an infected host.

As per estimates by our model, clearly; the Rn1 for top10 states listed below is greater than 1. This indicates that the infection rate is greater than the recovery rate, and thus the infection is currently in upward trajectory throughout India and the infection is expected to grow throughout the country.

**Figure 4:**
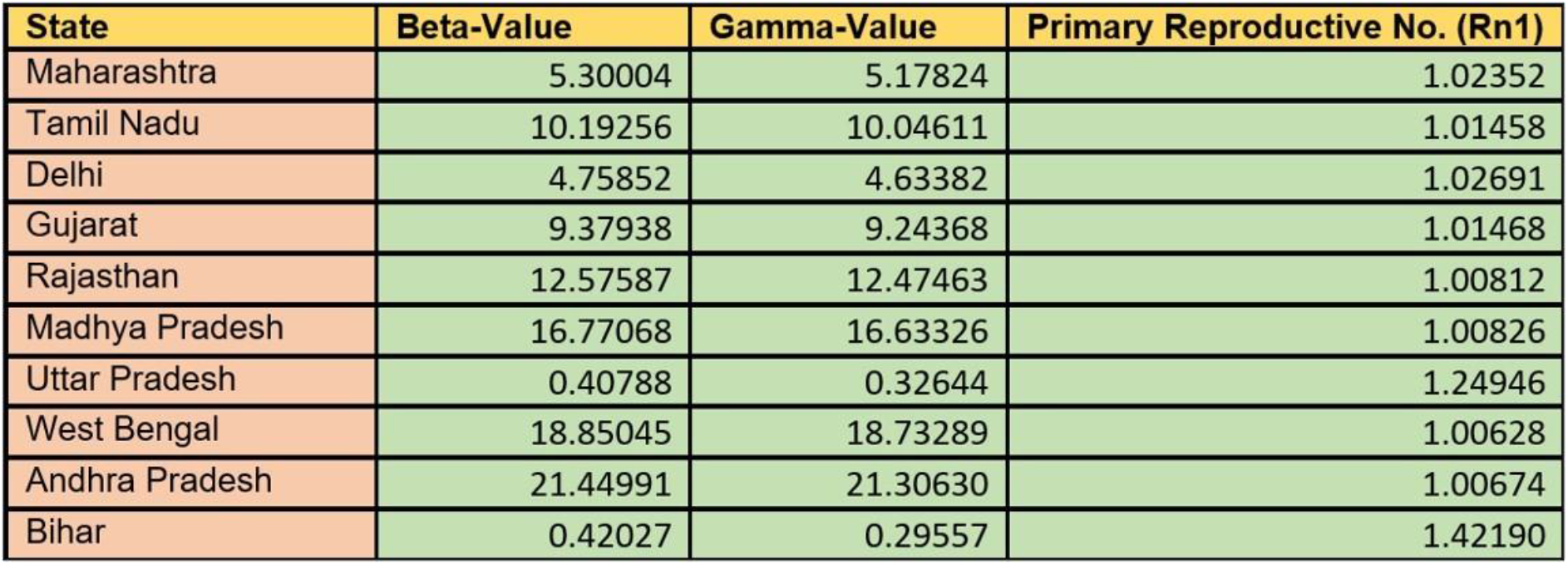
Estimated Values of reproductive number (Rn1) for top 10 affected states in India

Subsequently when we try to compare the expected peak for the top most states in India; the following observations can be deciphered. The predictive modelling was done using the SIR model parameters as the base model. Figure 5 predicts an estimation of case upsurge and peak estimation. These estimates depict that the most affected states such as Tamil Nadu, Rajasthan, Delhi and Gujarat are deviating by a delay of 5,4,6 and 11 days respectively in achieving infection peak in comparison to the SIR model prediction and the actual live data fetched from these states.

**Figure 5.**
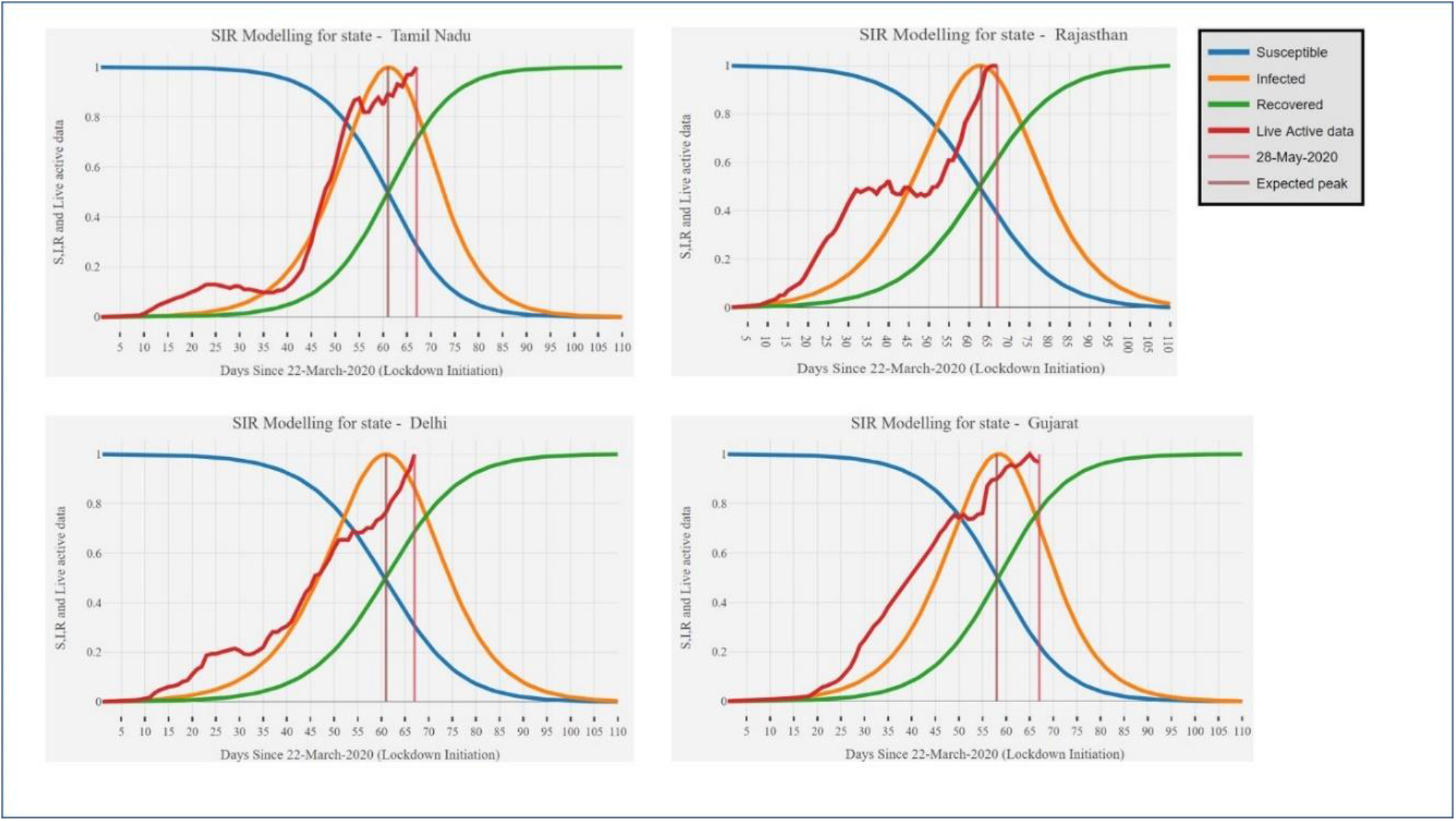
Peak estimation of affected states such as Tamil Nadu, Rajasthan, Delhi and Gujarat

In Phase 3 of our model, we included another significant parameter for Indian scenario vis-a vis mobility for SIR prediction; to estimate a new reproductive number. The dataset used for the construction and analysis of the graphs is provided in Supplementary file 1. Figure 6 depicts the mobility value, state population density, primary reproductive number (Rn1) and newly estimated secondary reproductive number (Rn2) for 10 most affected states. The value of secondary reproductive number was observed to be higher in most states except in Bihar, with Rn1 of 1.42 and Rn2 of 1.24. The current data is based on the Intra-state movements within the period of lockdown. The virus spread has been relatively lower for Bihar till 28^th^ May2020. This is most likely related to socioeconomic reasons related to lower number of individuals importing the virus from other COVID-19 affected countries in initial days. Accordingly, the status of disease spread in Bihar was low in the initial days. This can be corroborated with the primary reproductive number of Bihar which is low. However, the population of migrant labourers native to Bihar and working in metro cities which have high number of cases is around 3 million and with all these people returning to Bihar, the government have to be prepared for an upsurge because our data indicates highest mobility factor for Bihar. Low secondary reproductive number must be appropriated because of an initial low number of cases in Bihar; however this scenario may change due to higher number of migration to the state from other parts of the country.

**Figure 6.**
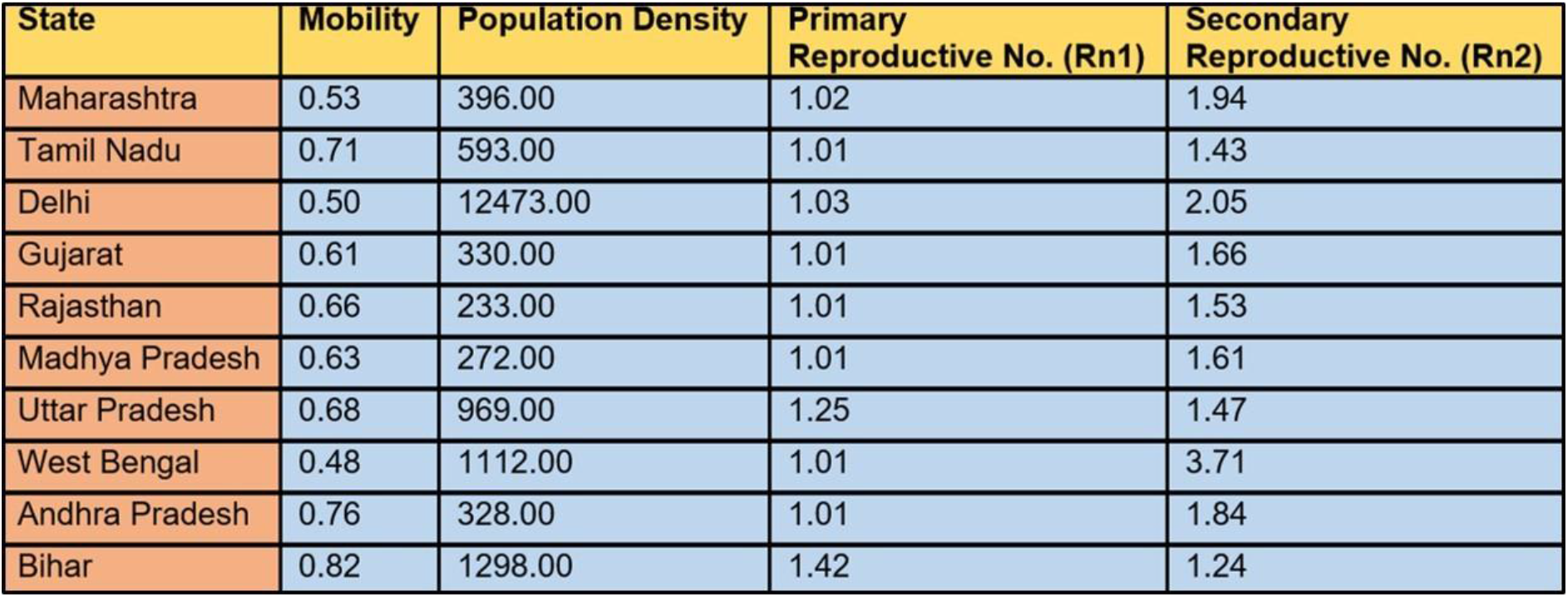
Estimation of secondary reproductive number (Rn2) for 10 most affected states

To determine relation between mobility, population density and reproductive number of each state, we performed correlation of these variables. In real world scenario, restrictions like lockdown change the mobility of the people and also compartmentalize the state in different zones. These restrictions decrease the correlation between population density and spread of disease. Further Lockdown measures are similar in all states but as population density varies in different region. Therefore we performed correlation test to determine if despite lockdown measures, there is any correlation between population density and Rn Figure 7 depicts the correlation matrix with correlation coefficient calculated using 3 different methods i.e. Kendall’s tau b Correlation, Pearson Correlation and Spearman’s rho Correlation.

Figure 7 shows a positive correlation between population density and Rn1 when data is assumed non-parametric (ranked), and shows positive correlation with Rn2 in all 3 methods. Estimates from our model indicate mobility is indirectly or negatively proportional to the populational density. This indicates that a state that is densely populated but under lockdown; is expected to have a lower number of cases. As the mobility in the state increases with more number of people going out from the highly affected states of the country to less affected states, whereas the states with a lower number of cases currently with the incoming flux of migrants and increase in the mobility, the number of cases will increase at higher rates. Negative correlation thus indicates that a decrease in population density is indicative of an increase in mobility. Since Rn2 was calculated by including mobility in the model, negative correlation represents the current situation in a state and impact of lockdown as explained in Figure 8. Also, since Rn1 does not account for mobility, it shows a positive correlation, suggesting that increase in mobility will increase the reproductive number of a state. A positive correlation was found between population density and both reproductive numbers as per estimates from our model. These predictive values obtained corroborate the fact that increase in population density and additionally due to influx of mobility factor in a state will increase the rate of COVID-19 infections. The correlation between mobility and Rn1 is statistically significant because calculation of Rn2 involves mobility itself. We found a positive correlation between mobility and primary reproductive number. Correlation was higher when data was assumed to be in normal distribution.

**Figure 7.**
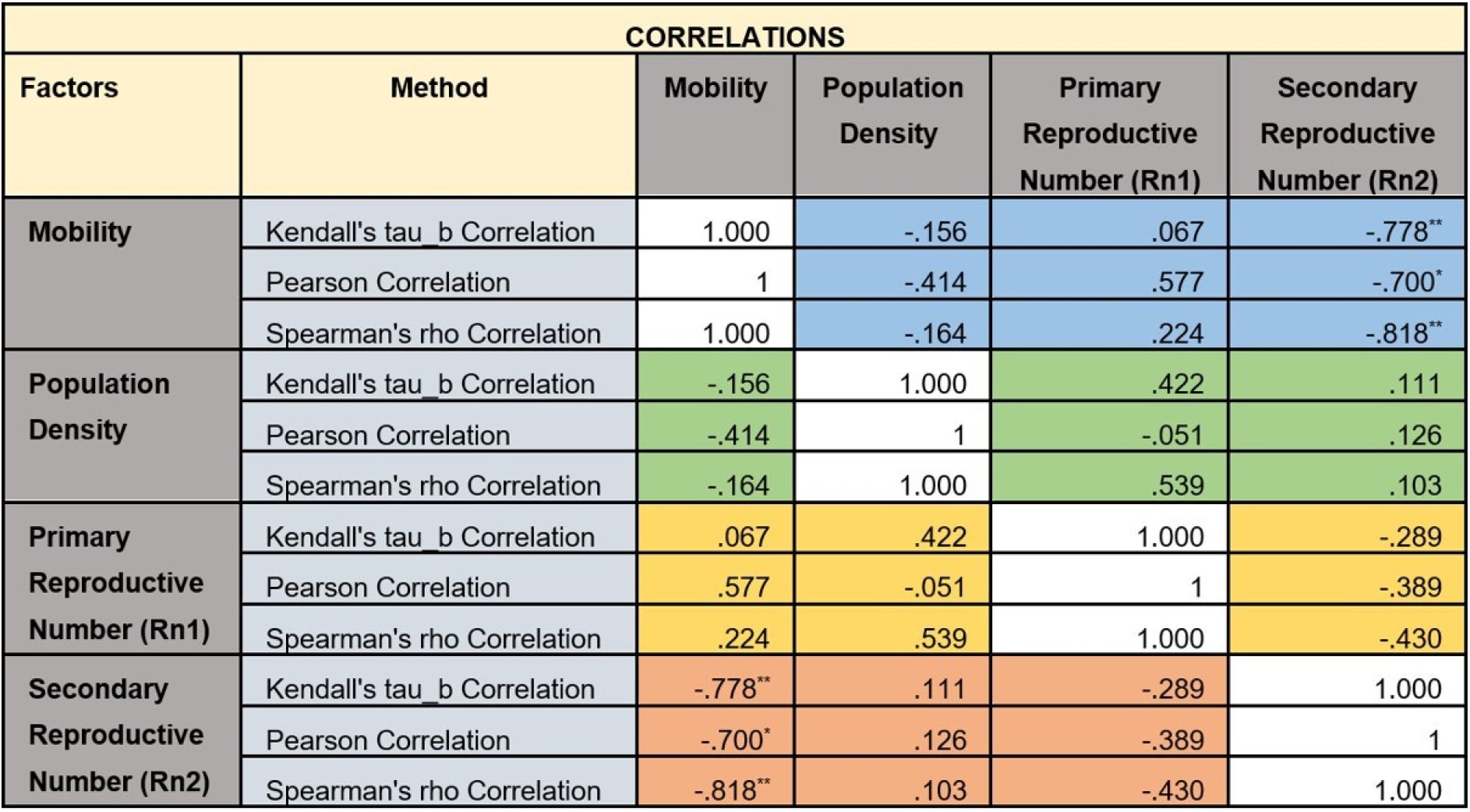
Correlation Analysis between mobility and population density

In the next phase we have tried to estimate the impact of mobility on the Reproductive number of 10 most affected states till 28^th^ May 20. Current situation in above mentioned states can be observed by scatter plot, which depicts the cause and effect result in a graphical manner for easy interpretation. Out these 10 states, R0 was observed to be highest for West Bengal with the lowest mobility number. This suggests that stricter lockdown measures are required in WB to control the spread of disease. Bihar on the other hand although has lowest reproductive number in the initial phase of the spread has been included in this graph as we have discussed before mobility is observed to be highest. This observation indicates that unless immediate intervention to curb mobility is done there is an imminent possibility of exponential rise in infection rates in Bihar.

**Figure 8.**
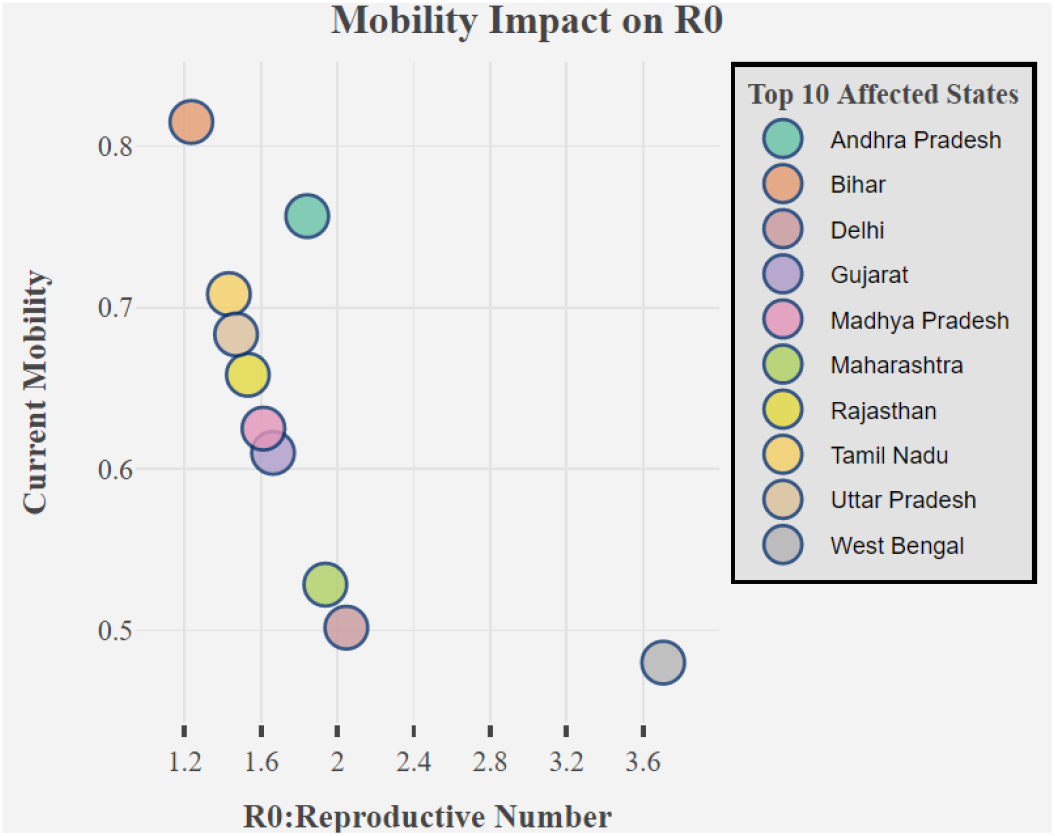
Impact of Mobility on Reproductive number of 10 most affected states

In phase 4 in our model, we mapped the current number of hospital beds with number of active cases present in a state. The dataset used for the construction and analysis of the graphs is provided in Supplementary file 2. The above deduction will help us to establish the preparedness of state in the light of outbreak. It has been established from literature that 80% of the COVID-19 affected cases in India are asymptomatic and do not require hospitalization [Onder et al, 2020]. As per the data available, India currently has approximately 1.9 million hospital beds, 95000 ICU beds, and 48000 ventilators. However, most of these beds and ventilators are concentrated in seven states – Uttar Pradesh (14.8%), Karnataka (13.8%), Maharashtra (12.2%), Tamil Nadu (8.1%), West Bengal (5.9%), Telangana (5.2%) and Kerala (5.2%). Delhi ironically is missing from the top states with regard to beds availability. Figure 9 depicts the availability of hospital beds in public hospitals, private hospitals and total beds in Maharashtra and Delhi as representative examples, as compared to the active cases in the above mentioned states respectively. The symptomatic cases can be assumed to be 20% of the active cases that require hospitalization and occupancy of hospital beds. Both these states have a high population density; however, Delhi has 39,455 hospital beds (govt and private) in comparison to Maharashtra which has 2,31,739 hospital beds (govt and private). However, there may be more number of asymptomatic cases in both Maharashtra and Delhi but as per the current scenario the data from hospital beds suggests that Maharashtra has sufficiently available hospital beds till date, whereas in Delhi, the data depicts lesser infrastructure. Therefore in comparison to Maharashtra and if the number of infected cases increases in Delhi; the demand of hospital beds will arise sooner in comparison to Maharashtra.

**Figure 9.**
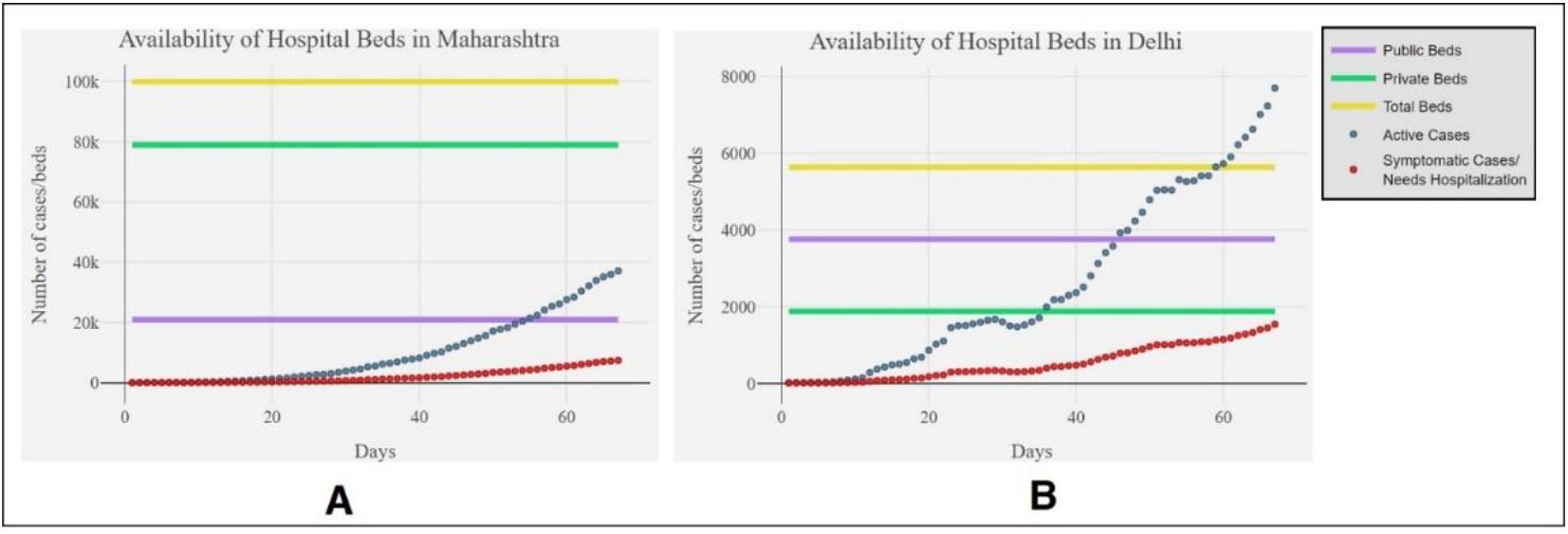
Comparative Data on Cases spike and availability of hospital beds in 2 major affected metro cities: Delhi and Maharashtra

Similarly, we compared currently available cases in Delhi and Maharashtra with the number of ICU beds and ventilators available in both states (Figure 10). For comparison we assumed demand of ICU beds and ventilators correlating with the number deceased cases. More explanation on this assumption can be found in Phase 4 of the model section later. Delhi has 1,973 ICU beds whereas with Maharashtra has 11,587 ICU beds. As depicted in Figure 9, the requirement of ICU beds and ventilators is more in Delhi than Maharashtra. Our model can be used to compare same for any other state or country.

**Figure 10.**
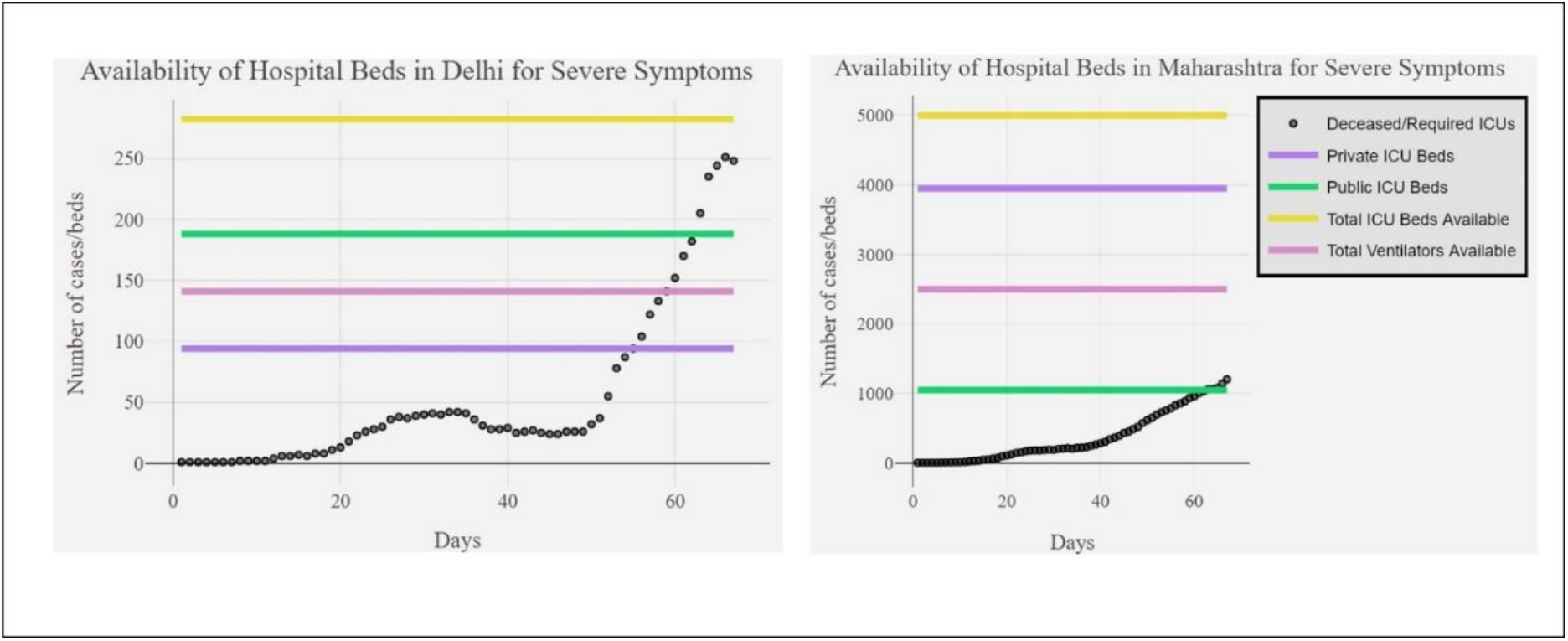
Correlation between cases in Delhi and Maharashtra with the number of ICU beds and ventilators available

Preventing disease outbreak and community transfer requires extensive testing and isolation of affected individuals. As the rate of epidemic increases, demand of testing laboratories with efficient testing methods also increases. In order to determine the preparedness of Indian states in this regard, we have compared the number of private, government and total laboratories available for testing of COVID-19 in the 10 most affected states in India (Figure 11). The dataset used for the construction and analysis of the graphs is provided in Supplementary file 3. Currently RT-PCR/TrueNat/CBNAAT and Antibody Rapid Test are the methods available for the testing [CDC, 2020]. Maharashtra has the highest number of total laboratories available, whereas least number of laboratories are available in Bihar. Since states in Figure 11, include top 10 most affected states, need of more testing laboratories is required in Bihar, Madhya Pradesh and Rajasthan as per our model. This will be especially significant for states such as Bihar that are expected to show an upsurge in infection due to migrant population mobility (Figure 7).

**Figure 11.**
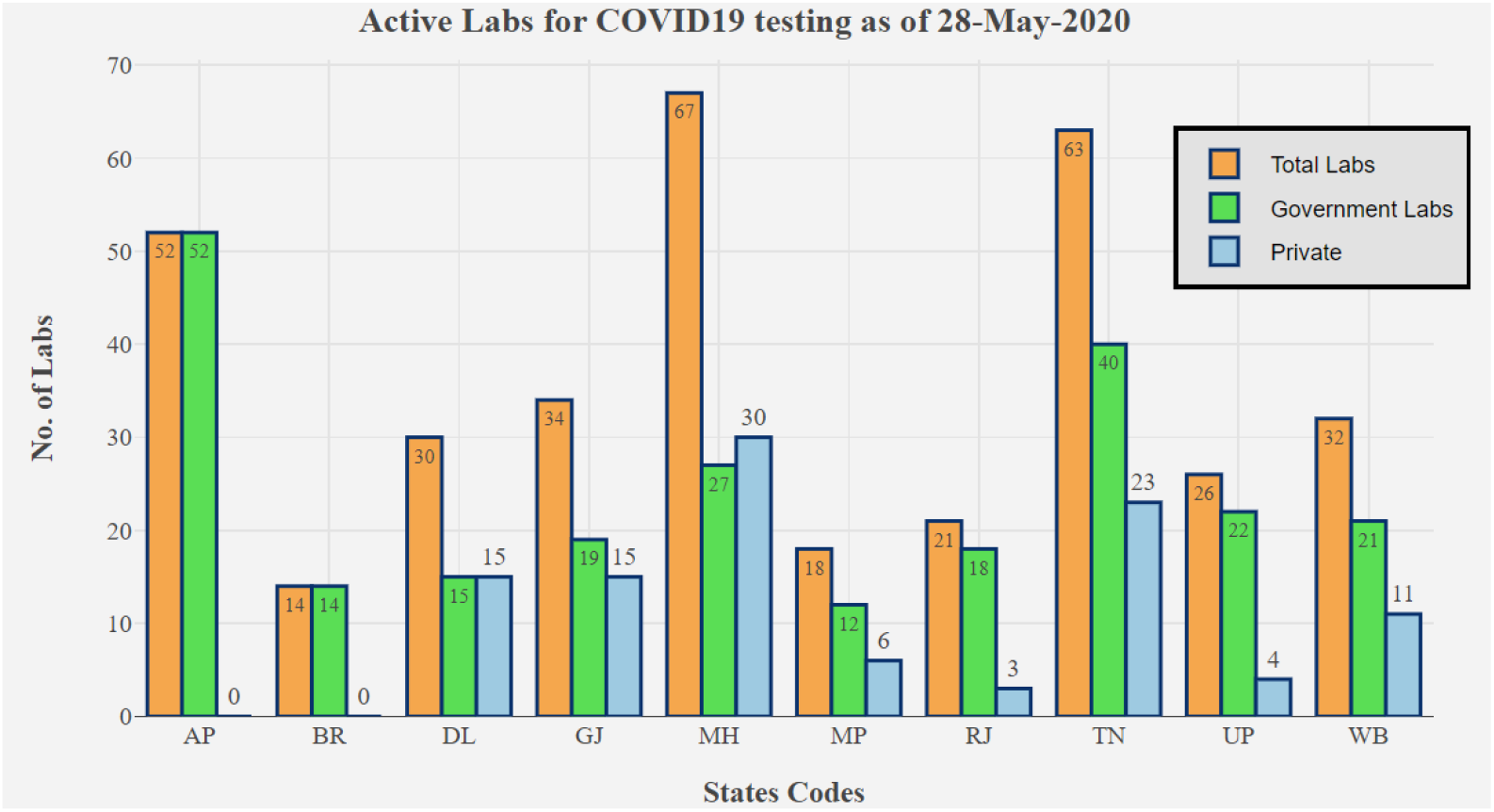
Representation of number of private, government and total laboratories available for testing of COVID-19 in the 10 most affected states.

Further, as on 28^th^ May 2020, ICMR has reported 34, 11,599 number of samples tested out of which 158,333 were found to be positive samples [ICMR, 2020]. Testing rate in an epidemic plays a crucial role in overcoming it. In SIR modelling, 100% cases are moved from susceptible to infective per unit time. However, in real life scenario diagnosis is required to report infective cases in a population. In order to curb an epidemic, testing rate should ideally be greater than susceptible decrement rate, but may not be replicated in real-time scenario. Higher the testing rate, higher is the probability of timely intervention for quarantine and treatment measures for infected individuals. Higher testing rate also decreases the exposure to susceptible individuals before the patient becomes symptomatic. Pool testing of the samples must be done on larger scales to cover maximum percentage of the stat population. Our model compares the current testing rate and active cases, with predicted cases to observe the requirement of increment in testing rate/day. In India, currently a laboratory performs an average of 150-200 tests/day [Bloomberg Quint, 2020]. Average rate is observed as the shaded green portion in Figure 12, which includes testing rate comparison for Gujarat, Madhya Pradesh and Delhi as representative examples. Prediction studies using our model indicate that an increased number of cases were identified when numbers of tests were intensified in states like Gujarat and Madhya Pradesh. However in case of Delhi a decline in number of cases can be observed. This interestingly is not due to downtrend in infections; rather it is the consequence of lesser number of tests being done in Delhi. Therefore, as per predictions of our model Delhi state requires to ramp up its testing infrastructure in order to contain the disease especially in the light of its population density.

**Figure 12.**
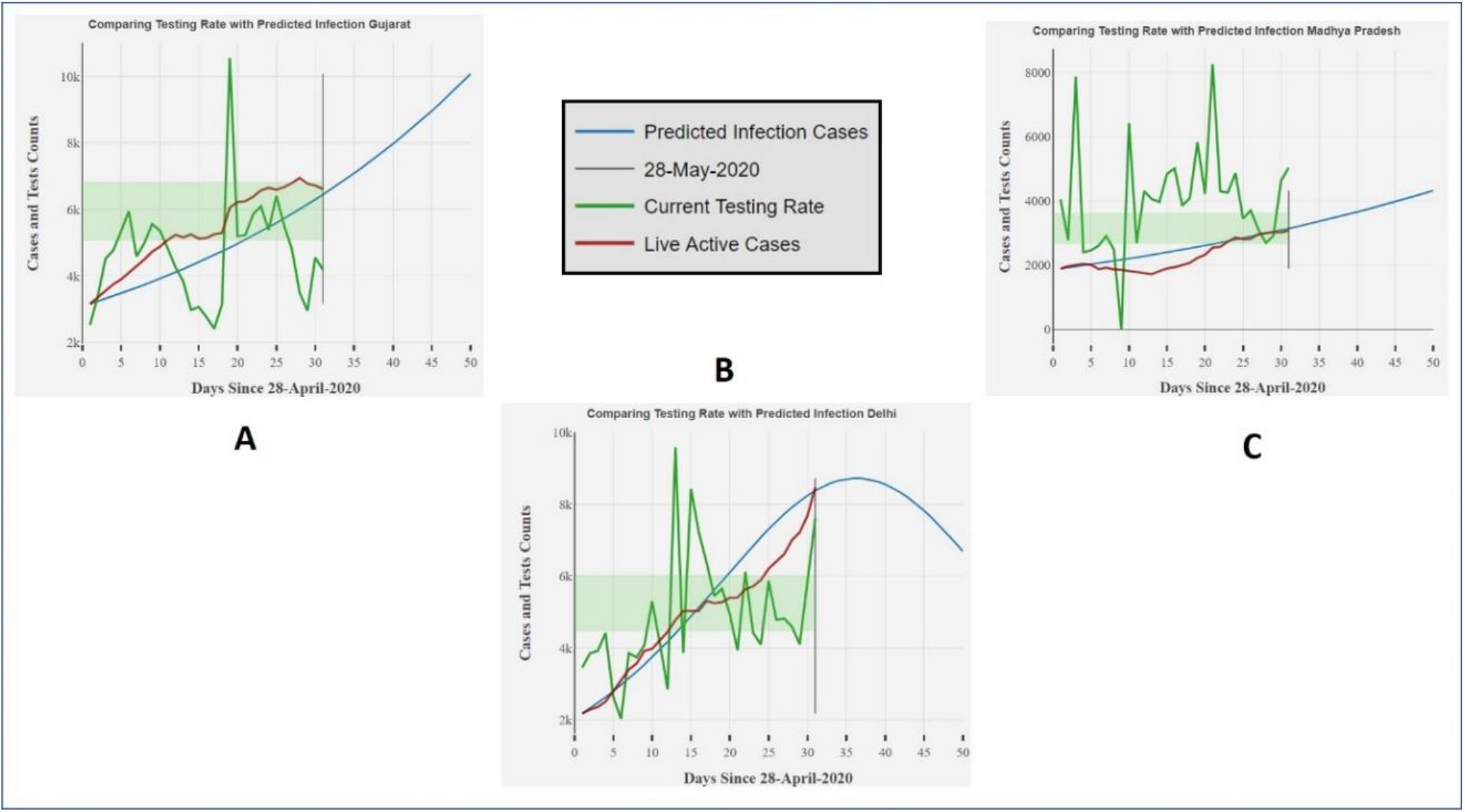
Correlation between predicted infection cases, testing rate and live active cases

## METHODS

In order to analyse epidemiology of COVID-19 and its impact in India, we built a modified SIR forecasting model to multiple datasets including infection numbers, mobility data, and hospital beds and testing ratios. The model was scripted using R programming language and is available on https://github.com/Samuel-Bharti/Epidimeology-Model-COVID-19. Inputs provided in the model are a collection of APIs (Application program interface) as well as offline dataset on secondary factors. Information like mobility, population area, etc participates in forecasting and correlations. Static graphical output is generated to visualize the impact of disease and its subsequent spread. An important consideration in our model is the ability to analyse state/district-level data rather focussing only at country-level. This provides a close relation in understanding of disease spread around an individual as well as, better prediction of trend in the smaller area. All the real-time data on number of cases are taken from 21^st^ March 2020 (api.COVID-19india.org) to analyse the effect after the lockdown.

## DATA SOURCES SELECTED FOR MODEL

The government sourced data are not often updated with same rate of changing statistics of epidemic. Hence, we additionally selected crowd source databases [API, 2020], which are more frequently updated to get the latest number on infection status and inpatient data, where patient information was stored from various validated reporting source. Government databases were used to validate crowd source data and to provide secondary inputs for our model like state population, state area [Statistics Times, 2020] and numbers of samples tested [API, 2020]. We used Google mobility reports [Google Mobility Reports, 2020] to get state-wise data on mobility reduction. They provide an estimation of reduction in mobility of individuals, at where categorical locations including office, parks, malls, transit stations, etc. Crowd source database [API, 2020] enabled us to investigate and analyse data on the state as well as district level. It also provided case-time series data for active, confirmed, deceased and recovered cases. Our model feeds on live datasets, by the use of API data to produce forecasting information and secondary effects. Availability of sufficient treatment components is extremely important in managing the widespread of disease. We used study done by Kapoor et al. 2020 to get dataset on available hospital beds, ICU beds and ventilators in each state of India [CDDEP, 2020]. In order to get accurate statistics on sample testing data we used health department and COVID-19 dashboard websites of every state and collected data from notice announcements published daily. All data and code of model required to reproduce the analysis is available online at https://github.com/Samuel-Bharti/Epidimeology-Model-COVID-19.

## MODEL CHARACTERISTICS AND PARAMETERS

### Phase 1: Descriptive Statistics

This phase utilizes the use of raw inpatient data fetched from crowd sourced database. It aims to extract gender and age statistics of the affected individual by cleaning the data. It categorizes age of the patient in 5-year range age groups defined in the model, starting from 1-5 years, 6-10 years, 11-15 years, and so on till a cap of age above 75. It produces donut pie chart for gender statistics, bar chart for number of cases in each age group and a bilateral bar chart of both gender and age statistics.

### Phase 2: SIR modelling and Coefficient Estimation

In our model, forecasting is implemented with SIR model along with the sum of squares and maximum likelihood method for coefficient estimation. SIR is the most used epidemiological model to study and forecast trends of a disease [Wangping et al. 2020]. Normal birth rate and death rate were assumed to be static. Ordinary differential equations calculated change in number of individuals in the 3 categories i.e. Susceptible, Infectious and Recovered. Beta value is the product of contact rate and transmission probability and taken as 0.35 [MAA, 2020] for default. Whereas, gamma value is 1/**infection** period, with infection period as an average of 5 days [Li et al. 2020], estimating gamma to 0.2 as default. The input data in SIR for S0 is the initial number from active cases of real time data. To improve analysis statistically and theoretically, real-time data for number of COVID-19 active, confirmed, deceased and death cases were taken. Sum of squares assume data in normal distribution whereas likelihood method assumed data to be in Poisson distribution. Beta and gamma are calculated from both methods, followed by optimization. Best fit of values are used for the forecasting of COVID-19 infection trend. Further, Rn1, reproductive number, defining number of secondary infections caused by an infectious individual, was calculated by the ratio of beta to gamma.

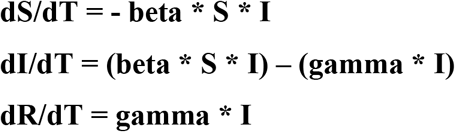

This phase produces estimation table with beta, gamma and Rn1 values along with SIR modelling graphs of each region mapped with live active cases for comparison.

### Phase 3: Secondary Factor Relations

Rn, reproductive number plays a key role in understanding and predicting spread of a disease. Phase 3 of the model shows the involvement and impact of secondary factors in the epidemiology of COVID-19. The secondary reproductive number (Rn2) estimated in this phase, is based on the condition that lock down is already imposed and measures are being taken to decrease and stop the widespread of COVID-19. Factors included are mobility reduction in each state since lockdown and state population density. The percentage of average reduction in mobility is converted into a numerical coefficient in model. We introduced a lock down coefficient in SIR, Rho, consisting of two data inputs, mobility pattern average and ratio of essential services to total services in a city. Apart from these two factors our model is capable of including an additional data input on service status, which reports the total number of essential services in a state during epidemic, divided by total services in a state without epidemic.

Lockdown Coefficient is defined as below:

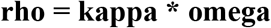

where kappa is the mobility,

omega is the ratio of essential state services to the total state services

Here services status was ignored due to data unavailability, and coefficient kappa was equated to rho. Rho ranges from 0 to 1, where 1 meaning no lockdown and 0 being complete lockdown and shutdown of all individual interaction. Introducing Rho will affect the equations as follows:

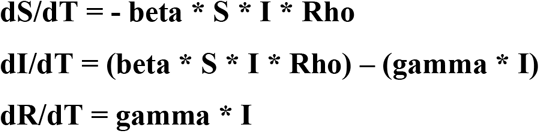

Assuming individual as the gas molecules, state as the container, and mobility as the interaction/collision resulting in spread of infection. According to kinetics of gas molecule, the collision probability should be dependent to number of molecules and the container area. In other words, collision is depending upon molecule density; similarly we can equate interaction/spread with state population density. To analyse the relation, we correlated population density with estimated reproductive numbers and mobility. This phase produces correlation matrix table using Kendall’s tau b Correlation, Pearson Correlation and Spearman’s rho Correlation to correlate mobility coefficient, population density, primary reproductive number (Rn1) and secondary reproductive number (Rn2).

### Phase 4: Treatment Components Relation

Model compares the number of active cases with available hospital beds in the region and number of deceased cases with available ICU beds and ventilators, by assuming deceased cases had depicted severe symptoms and need for same. For this study, we additionally accounted for symptomatic and asymptomatic cases with the need of hospitalization and requirement of hospital beds by symptomatic cases. To compare deceased cases with availability of ICUs and ventilators, model generates a new dataset assuming a deceased individual using an ICU or ventilator average of 15 days before death. Hence, occupancy of each ICU or ventilator is reset in every 15 days. Equation for same is as below:

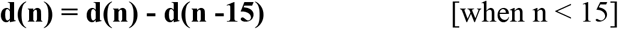

Where d is the number of deceased cases or occupied ICU bed/Ventilator and n is the number of days.

### Phase 5: Samples Testing Data Statistics and Predictive Impact

Phase 5 emphasize on the analysis of sample testing data. It plots a bar graph between the number of laboratories available and their type for each state, to visualize the need of more laboratories in a region according to the number of cases and reproductive number. Additionally, this phase accounts the testing rate of each region and compares it with active cases in the region as well as with the predicted cases. It also plots the current average testing rate as the shaded region. Aim of this analysis is to determine the efficiency of current testing rate. Ideally that rate of sample testing should be greater than the rate of decreasing of susceptible individuals.

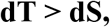

where dT is current testing rate and dS is the predicted susceptible decrement rate.

This comparison is independent of the outcome of test, whether individual is found to be positive or negative, rather it highlights the demand of more laboratories to test every individual timely, which will contribute in decreasing reproductive number and rapid decline in new infective cases.

## Discussion

Through our model, we have taken inpatient data, state population and density data, mobility data, hospital beds, ICU availability and testing data as input parameters. With these input parameters, we can predict and perform several analyses for this pandemic. In the first part of the model, we predict the descriptive parameters such as statistics of male-female cases effected in a state/country; and provides age-wise cases effected in a state/country. In the second part of the model, we present our version of the classical SIR model, and estimate beta and gamma according to present/active data of COVID-19 cases. Through this we were predict the R1 and future trends of COVID-19 outbreak for each state. In the third phase, we have compared the state population density with R1 and calculate the lockdown variable using mobility data and services status in a state. In the fourth phase, we have compared the treatment components relation where we predict the requirement of beds, according to predicted number of cases. In the last and final phase of our model, we predict testing rate according to current number of labs present and tests done daily and compare it to reproductive number (Rn1) to predict if people are detected on time or not.

Our model is instrumental in establishing links between important parameters such as population density and mobility for predicting the upsurge of the cases in the coming times. The model exhibits an exquisite feature of application program interface (API) dataset connectivity as the input for time dependent data. The model is thus useful to generate the graphs as per the changing input parameters and provides a dynamic solution to the prediction of disease spread with the ever-growing numbers. Our model highlights and quantifies factors required to efficiently resolve the current pandemic. The model also serves as the dynamic solution by featuring the option of API dataset connectivity as the input for time dependent data. The model thus calculates required coefficients and generates output depicting future trend and requirements as per current situation. It can explain and predict long-term effects with different lockdown driven variables. The 5-phase in-depth analysis helps in understanding and predicting the dynamic evolution of the pandemic, based on current statistics. The model focusses on estimation and prediction, by evaluating current inputs to sufficiently handling the pandemic such as, the need of testing centre or testing kits can be predicted using our model as well as can be validated for sufficiency according to upcoming number of cases. The infective case predictions are not sensitive for exact number of cases but provide estimation data to correlate with number of beds, testing centres, testing capacity and other responsive factors.

Along with mobility, the model also considers the correlation with population density, which theoretically plays an important role in the evolution of pandemics. Though, the correlation results can vary due to the efficiency of lock down measures and implementation in each state/country. Absence or failure of lockdown during the pandemic, will show high positive relation between reproductive number and population density. The low or negative correlation between density and Rn1 shows the compartmentalization in the region which is a positive impact of efficient lockdown measure. Other variable can be included in the lockdown driven coefficient, depending upon availability of data, but mobility data can be assumed to substitute as a total.

Asymptomatic case time data have been ignored in the model due to unavailability, but an approximation of 80% asymptomatic cases was used in phase 4 results. Phase 5 in the model substitutes the need of testing portals and efficiency. Also, in viral infections, the virus is most likely to mutate, causing different biological effect which may be symptomatic later, if not earlier. So, it is necessary to primarily focus on treatment infrastructures and their establishments at the earliest during the pandemic. The model can be used to estimate impact evolution of epidemic caused by other diseases.

## Conclusion

The outbreak of COVID-19 has been labelled as a black swan event [Wind et al.2020]. This pandemic has had a detrimental effect on global healthcare systems with a ripple effect on every aspect of human life. Therefore, predictive modelling can be an important tool to minimize both economic and community impact. This is especially important for a country like India that has several limitations with regard to healthcare infrastructure, diversity in socioeconomic status, high population density, housing conditions, health care coverage that can be important determinants for the overall impact of the pandemic. The results of our 5-phase model depict a projection of the state wise infections/disease over time. Our model can generate live graphs as per the change in the data values as the values are automatically being fetched from the crowdsourced database. Asymptomatic cases have been ignored in the model due to unavailability of such data on public platforms. Also, phase 5 analysis in the model indicates the need of more testing portals and increased efficiency. In future, we intend to build a web-interface for generation of graphs to make it user-friendly and easy to understand with inclusion of more parameters as in the Indian context.

## Data Availability

All the methodology and supporting data is provided in supplementary files for the reproduction of the results. Link for code: https://github.com/Samuel-Bharti/Epidimeology-Model-COVID-19. For gender, age and number of cases in each state, data was dynamic. Main source: https://api.COVID-19india.org/

## Supporting Information

- Supplementary 1: An excel sheet of value of mobility across states
- Supplementary 2: An excel sheet of state parameters oh hospital beds, population, area, density and ICU beds
- Supplementary 3: A list of reactions used with abbreviations and equations. (PDF)

## Acknowledgements

We would like to acknowledge Dr. Ashok K. Chauhan, Founder President, Amity University Uttar Pradesh for providing us the opportunity to conduct research. We would also like thank Centre for Computational Biology and Bioinformatics and Centre of Medical Biotechnology, Amity Institute of Biotechnology, Amity University for providing us necessary resources.

## Author Information

### Affiliations

Amity Institute of Biotechnology, Amity University, Uttar Pradesh, INDIA.

